# Emergence of evidence during disease outbreaks: lessons learnt from the Zika virus outbreak

**DOI:** 10.1101/2020.03.16.20036806

**Authors:** Michel J Counotte, Kaspar W Meili, Nicola Low

## Abstract

**Introduction:** Outbreaks of infectious diseases trigger an increase in scientific research and output. Early in outbreaks, evidence is scarce, but it accumulates rapidly. We are continuously facing new disease outbreaks, including the new coronavirus (SARS-nCoV-2) in December 2019.The objective of this study was to describe the accumulation of evidence during the 2013-2016 Zika virus (ZIKV) outbreak in the Pacific and the Americas related to aetiological causal questions about congenital abnormalities and Guillain-Barré syndrome.

**Methods:** We hypothesised that the temporal sequence would follow a pre-specified order, according to study design. We assessed 1) how long it takes before findings from a specific study design appear, 2) how publication of preprints could reduce the time to publication and 3) how time to publication evolves over time.

**Results:** We included 346 publications published between March 6, 2014 and January 1, 2019. In the 2013-–2016 ZIKV outbreak, case reports, case series and basic research studies were published first. Case-control and cohort studies appeared between 400–700 days after ZIKV was first detected in the region of the study origin. Delay due to the publication process were lowest at the beginning of the outbreak. Only 4.6% of the publications was available as preprints.

**Discussion:** The accumulation of evidence over time in new causal problems generally followed a hierarchy. Preprints reduced the delay to initial publication. Our methods can be applied to new emerging infectious diseases.

## 1 Introduction

Outbreaks of infectious diseases trigger an increase in scientific research and output. Early in outbreaks, evidence is scarce, however, it accumulates rapidly as time progresses. As we are continuously facing new disease outbreaks, as we do at the moment with the emergence of the new coronavirus (SARS-nCoV-2), understanding the accumulation of evidence is vital. Here, we describe the accumulation of evidence during the Zika virus outbreak and summarize the lessons learnt.

Causality is a principal theme in epidemiological research. Establishing that an exposure causes a specific health outcome is based on evidence and may inform guidance about public health measures. The concepts and types of evidence required to conclude that an association is causal are the subject of ongoing debate. Vandenbroucke proposed a hierarchy of evidence based on the best chance for discovery and explanation of phenomena [1]. Observations published in case reports and case series, or findings in data and literature drive discovery. Verification of these discoveries happens in observational studies and in randomized controlled trials, given that exposures can be randomized. The value of evidence used for public health guidance is thought to follow an inverse pattern of the hierarchy for discovery; here, case reports and other anecdotal evidence is considered to be evidence that provides the least certainty on an effect or association. Cohort studies, randomized controlled trials and case-control studies are considered to provide evidence with a higher certainty (Figure 1).

**Figure 1.**
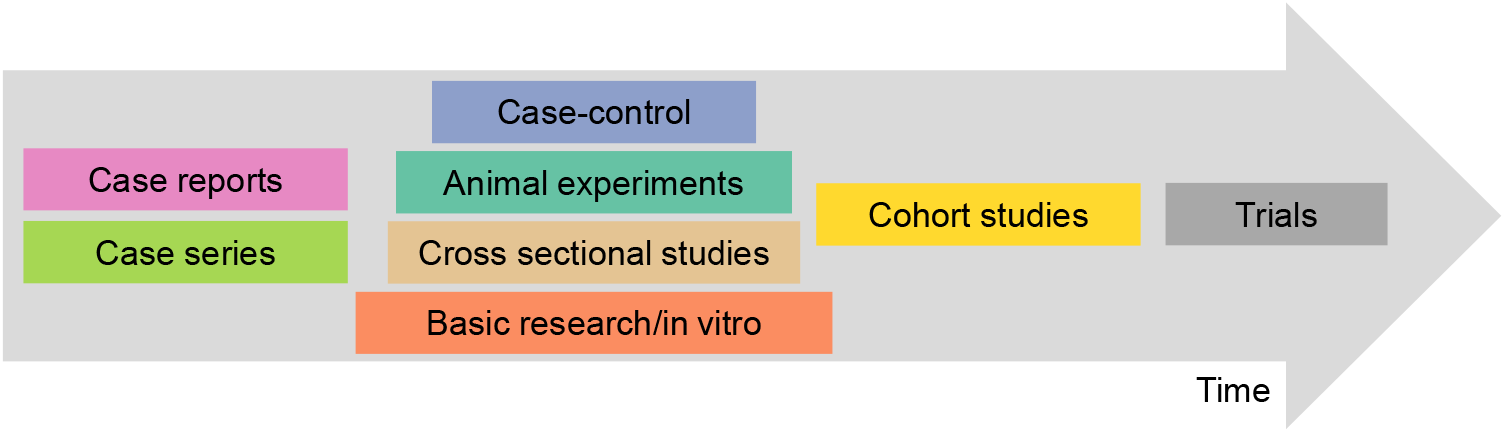
Hypothetical accumulation of evidence over time, by study design.

The Zika virus (ZIKV) outbreak in the Pacific and the Americas between 2013-–2016 presented several aetiological causal questions. In 2013—2014, ZIKV caused an outbreak in French Polynesia [2, 3]. During this period, investigators documented some severe neurological conditions, including 40 people with Guillain-Barré Syndrome (GBS). GBS is usually a rare sporadic condition. Often triggered by infection, an autoimmune response affects the peripheral nerves, leading ascending paralysis, which can be fatal if it involves the respiratory nerves [4]. At the time, the reports did not attract much attention and the investigators refrained from making a causal connection because dengue was also circulating at the time [3]. In November 2015, the ministry of health in Brazil reported a cluster of births affected by microcephaly in the north east. Microcephaly is a birth defect, indicative of impaired brain development, which can be caused by congenital infection. At around the same time, ZIKV had been identified for the first time in Brazil [5]. In December 2015, the Pan American Health Organization announced heightened surveillance owing to an “increase of congenital anomalies, Guillain-Barré syndrome, and other neurological and autoimmune syndromes in areas where Zika virus is circulating” [6]. The World Health Organization (WHO) declared a Public Health Emergency of International Concern (PHEIC) on 1 February 2016 because of the severity of these clinical conditions and their temporal association with ZIKV circulation [7]. Retrospective assessment of the French Polynesia outbreak identified an increase in adverse congenital outcomes as well [8]. The PHEIC and the extensive outbreak catalysed the research on ZIKV. Early public health guidance about the prevention of ZIKV infection and its potential consequences was based on limited evidence, however [9].

Systematic reviews were developed to address the PHEIC recommendation for research about the causal relationships between ZIKV infection and adverse congenital outcomes, including microcephaly and between ZIKV and autoimmune outcomes, including GBS [10]. The reviews organized the findings around a ‘causality framework’ with ten dimensions derived from those proposed by Bradford Hill [10, 11]. An expert committee reviewed the evidence collected by these systematic reviews up to May 2016 and reached the conclusion that “the most likely explanation of the available evidence” was that ZIKV is a cause of adverse congenital outcomes and a trigger of GBS [12]. This review has been kept up to date as a living systematic review, by periodically incorporating new results [13, 14]. The additional evidence has reinforced the conclusions of causality.

A temporal sequence for the emergence of evidence was already hypothesised during the planning of the systematic reviews in early 2016 (Figure 1). Acknowledging that ‘astute observations’ of new causes of disease often start an aetiological investigation [15], case reports and case series were eligible for inclusion in the systematic reviews. These study designs are often excluded from systematic reviews because they are the lowest level of the “hierarchy of evidence” that applies to evaluation research.

Vandenbroucke proposed a reverse hierarchy for discovery in which ‘anecdotal’ forms of evidence are at the top [1]. Cross-sectional, case-control and retrospective follow-up studies follow because they are quickest study designs that include a control group. Prospective cohort studies take longer to set up and RCTs only provide additional information if a treatment or vaccine is available. In addition to epidemiological studies, basic and clinical laboratory science start early in the search for causes.

The objective of this study was to examine the body of evidence that was used to establish the causal relation between ZIKV infection and adverse outcomes. We hypothesised that the temporal sequence would follow Figure 1.

## 2 Methods

### 2.1 Included studies/Review methods

We analysed records of studies that were included in published systematic reviews [10] and two updates [13, 14] of the relationships between ZIKV and congenital abnormalities and GBS. The methods of the original review and updates are described elsewhere [10, 13]. The included studies reported evidence about any of the questions of the causality framework, based on the Bradford Hill dimensions of causality. Until January 18, 2017, we included epidemiological and basic research study designs; after that date we continued the review of evidence from epidemiological study designs only (Figure 2). Here we analyse studies collected until January 1, 2019.

**Figure 2.**
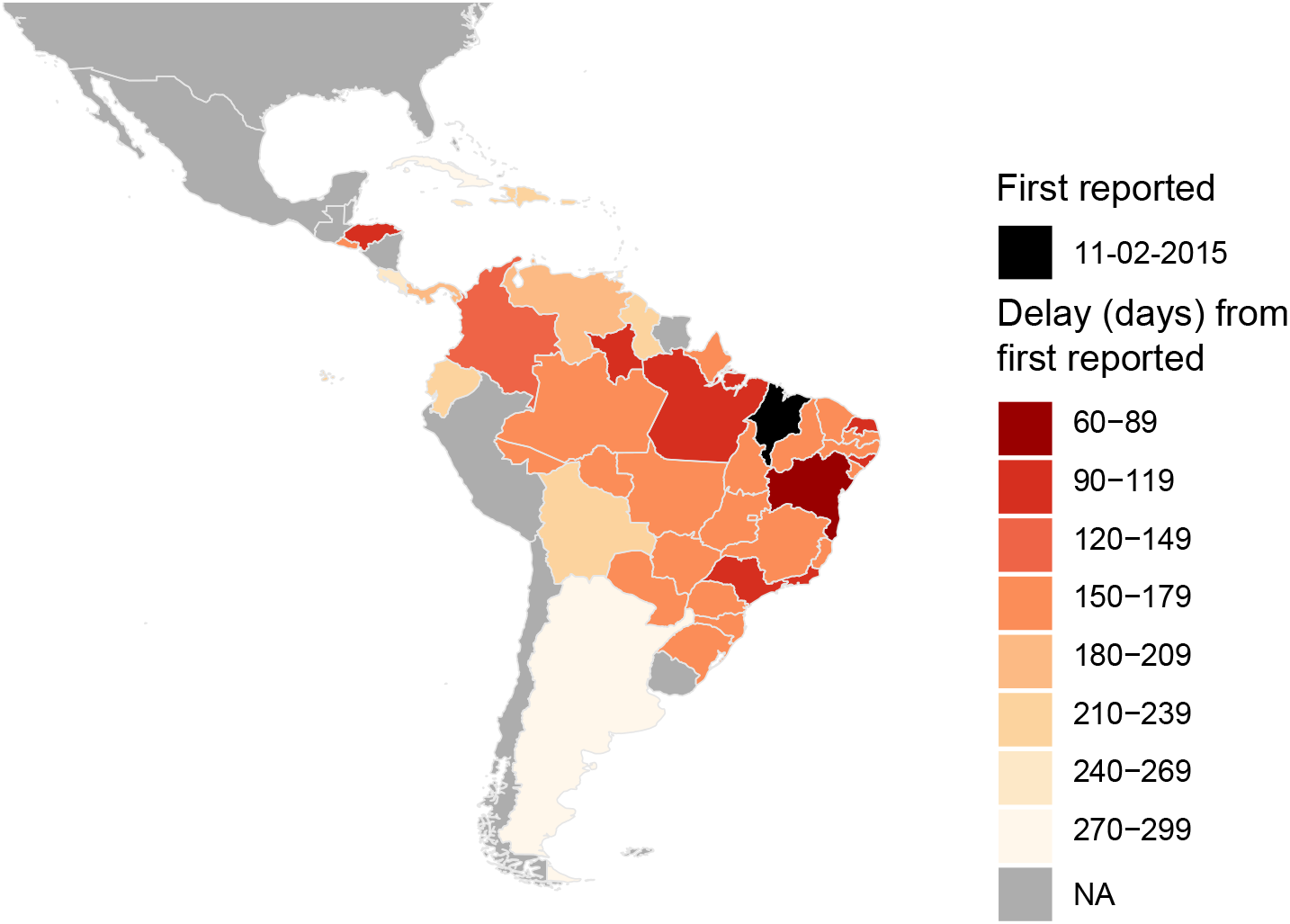
Map of South and Central America showing the timing of the first reported case of Zika virus according to the Pan-American Health Organization [16] by state for Brazil, by country for all other regions. NA: Not available.

### 2.2 Extracted information

For all included studies, we retrieved the received and published date, the location of the study and the study design (Table 1). For epidemiological studies, we extracted the study location and the number of patients with both exposure and the outcome according to the case definition provided in the publication. We excluded modelling studies or surveillance and outbreak reports.

**Table 1.** Information used in the analyses: Variables retrieved from PAHO, extracted from included studies (publications), and variables calculated from the data. Abbreviations: PAHO, Pan American Health Organization; PHEIC, Public Health Emergency of International Concern; ZIKV, Zika virus.

| Variables | Explanation | Source |
| --- | --- | --- |
| Introduction date | Date the state (for Brazil) or country reported the first case or cases of ZIKV according to PAHO [16] | PAHO |
| Publication date | The first date a publication was available | Publication |
| Received date | The date the publication was received by the journal the publication appeared in | Publication |
| Accepted date | The date the publication was accepted for publication by the journal the publication appeared in | Publication |
| Study design | Epidemiological studies: Case report, case series, case-control study, cohort study; Basic research: *in vivo* or *in vitro* studies | Publication |
| Outcome | Guillain-Barré syndrome or congenital abnormalities | Publication |
| Sample size (N) | For epidemiological studies: The total number of patients with the outcome and positive ZIKV exposure according to criteria of publication | Publication |
| Total time to publication from introduction | The time between introduction date and publication date in days | Calculated |
| Total time to publication from PHEIC | The time between February 1, 2016 and the publication date in days | Calculated |
| Publication delay | The time between the received date and the publication date in days | Calculated |

### 2.3 Introduction date

We considered the date the first case of endemic ZIKV was reported in each state for Brazil, or country for the rest of the region (Figure 2) [16]. We assigned 16 July, 2015 as the date of ZIKV introduction if the state in Brazil was not explicitly reported. We assigned 1 October 2013 as the introduction date for French Polynesia [17].

### 2.4 Publication date

We defined the publication date as the earliest date the publication was available. If the publisher’s website did not state an exact date, we assigned the ‘epub’ date from MEDLINE via PubMed or ‘page created’ date for specific online journals (EID and MMWR). We also recorded the date the manuscript was received by the publisher (received date) and the date of acceptance for publication (accepted date).

### 2.5 Total time to publication

We defined the time to publication as the time between introduction of ZIKV virus in the region and the publication date. For basic research studies, many of which were done in countries unaffected by ZIKV, we assigned the time to publication as the time between 1 February 2016 (the PHEIC declaration) and the first available publication date.

### 2.6 Publication delay

The delay resulting from the publication process (publication delay) was defined as the time between the ‘received date’ and the first available publication date.

### 2.7 Analysis

We assessed 1) how long it takes before findings from a specific study design appear, 2) how publication of preprints could reduce the time to publication and 3) how time to publication evolves over time. We provide a descriptive analysis of the total time to publication and the publication delay by publication. Of these durations, we provide the median and interquartile range (IQR) by study design and over time. We compare the publication delay by three month period (quarter).

## 3 Results

During the period of the first review [10] and subsequent update [13], we screened 2,847 publications. During the remaining period, between January 7, 2017 and January 1, 2019, we screened an additional 2,594 publications. Figure 3 shows the evolution of the volume of the published ZIKV research over time is provided.

**Figure 3.**
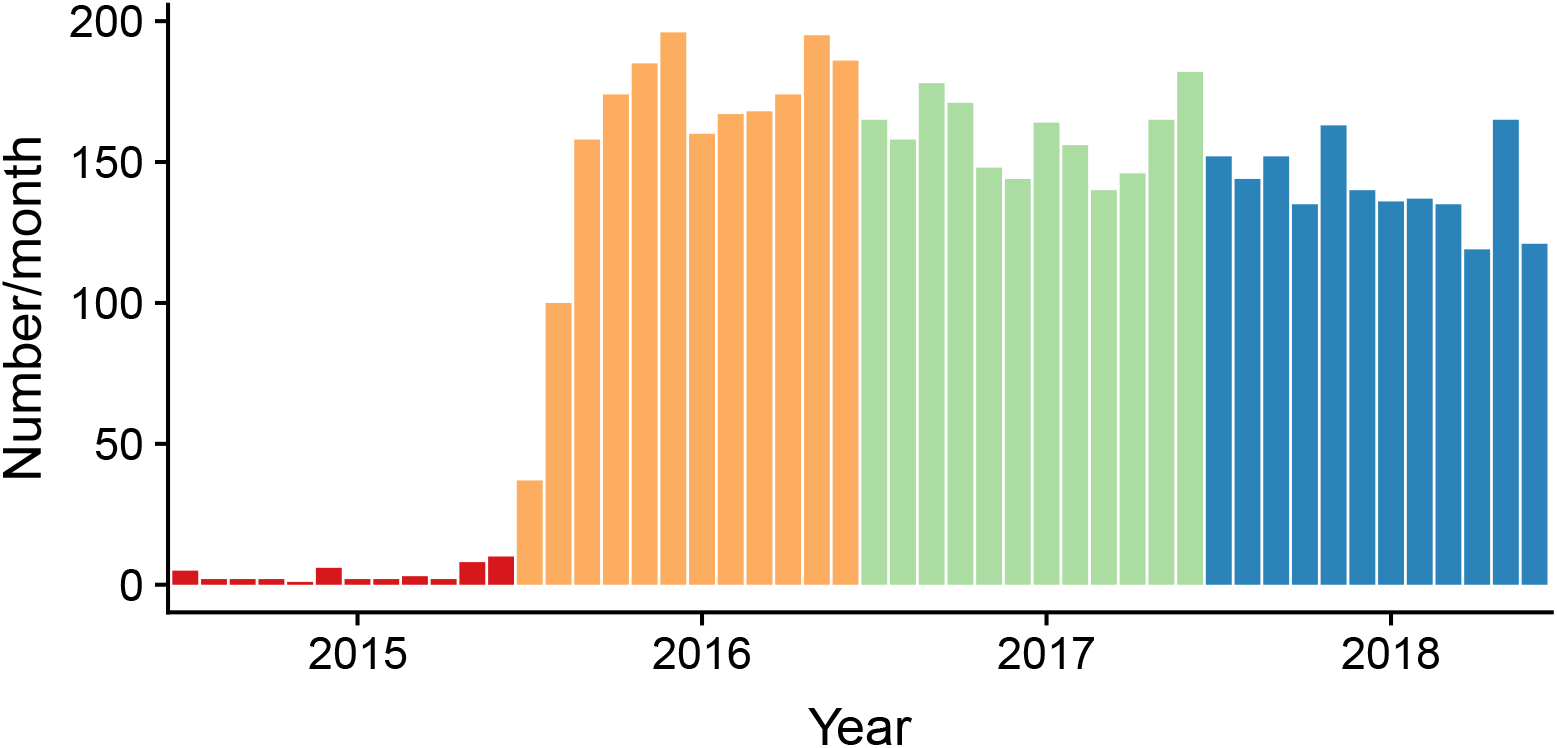
Research volume by month between 2016 and 2019, retrieved from MEDLINE via PubMed using the keywords ‘Zika’ and ‘Zika virus’.

### 3.1 Included studies

We included 346 publications published between March 6, 2014 and January 1, 2019 (Table 2 and Figure 4). Up to January 18, 2017, we included 171 publications. Most publications were epidemiological study design (94/171), 77 out of 171 studies were basic research studies (either research on animal models or *in vitro* laboratory-based research). Between January 18, 2017 and January 1, 2019, we restricted our search to publications of epidemiological study design, and included another 175 publications. For 220/269 epidemiological studies, a date of ZIKV introduction was known. The time between received and published was reported in 204 out of 346 studies. In 16/204 of these studies there was no publication delay, usually because the first publication date was as a preprint.

**Table 2.** Overview of counts and completeness of data per study design and outcome. Studies published up to January 1, 2019 for epidemiological studies reporting on at least one individual with ZIKV exposure and outcome of interest, and up to January 17, 2017 for basic research studies. Abbreviations: CA, congenital abormalities; GBS, Guillain-Barré syndrome.

| Study design/parameter | CA | GBS | Total |
| --- | --- | --- | --- |
| **Epidemiological studies** |  |  |  |
| Case report | 54 | 28 | 82 |
| Case series | 85 | 30 | 115 |
| Case-control study | 10 | 9 | 19 |
| Cohort study | 35 | 0 | 35 |
| Cross-sectional study | 14 | 4 | 18 |
| **Total epidemiological studies:** | 198 | 71 | 269 |
| Introduction date available | 159/198 | 61/71 | 220/269 |
| **Basic research studies** |  |  |  |
| Animal experiments | 24 | 0 | 24 |
| *In vitro* experiment/other | 51 | 2 | 53 |
| **Total basic research:** | 75 | 2 | 77 |
| **Total:** | 273 | 73 | 346 |
| Publication delay available | 167/273 | 37/73 | 204/346 |
| Studies with a zero publication delay | 16/167 | 0/37 | 16/204 |

**Figure 4.**
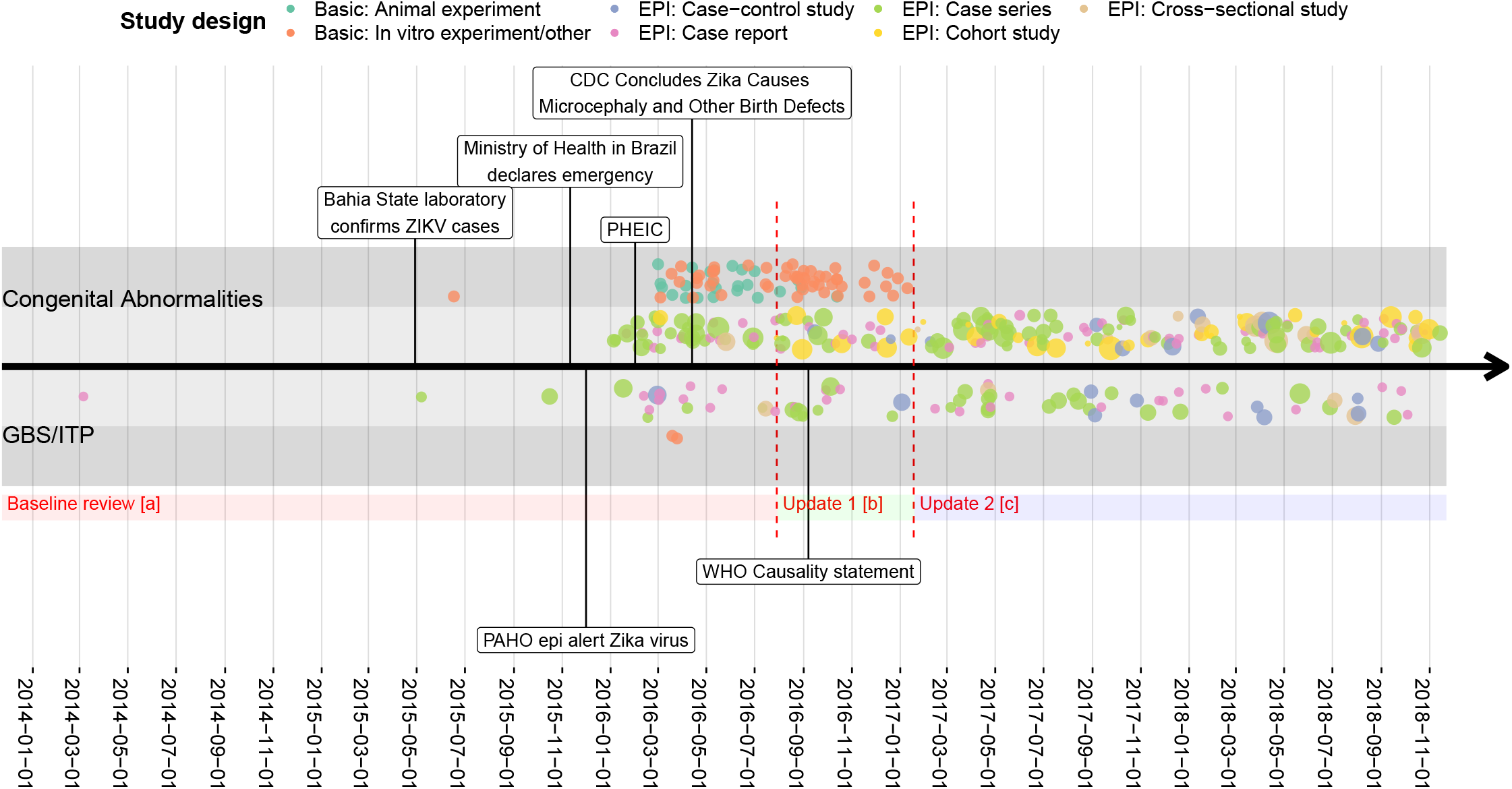
Publications by publication date, study design and outcome. For epidemiological studies, the size of the points correspond with the number of individuals with a positive outcome (Congenital abnormalities or GBS/ITP) and a positive exposure (Zika virus infection). Basic research studies were included up to the end of the first update (red dotted line Abbreviations: EPI, of epidemiological study design; CDC, Centers for Disease Control and Prevention GBS, Guillain-Barré syndrome; ITP, Immune thrombocytopenic purpura; PAHO, Pan American Health Organization; WHO, World Health Organization;, PHEIC, Public Health Emergency of International Concern; ZIKV, Zika virus. Different review periods: Baseline review [a] [10], update 1 [b] [13], update 2 [c] [14].

### 3.2 Total time to publication

Figure 5A shows the comparison of publications published between the PHEIC and the end of the second review period (January 18, 2017). We saw the first case reports and case series published after 44 and 77 days, respectively. Basic research emerged rapidly after the PHEIC. In this period, a limited number of case-control studies was available. The earliest publication of a case-control study for GBS, was a result of a retrospective study looking back at the French Polynesia outbreak [8]. The median total time to publication was longer for more robust study designs (cohort studies, case-control studies). We see a similar pattern for epidemiological studies if we consider the data up to January 1, 2019 and consider the time to publication between the regional introduction of ZIKV and the publication date (Figure 5B).

**Figure 5.**
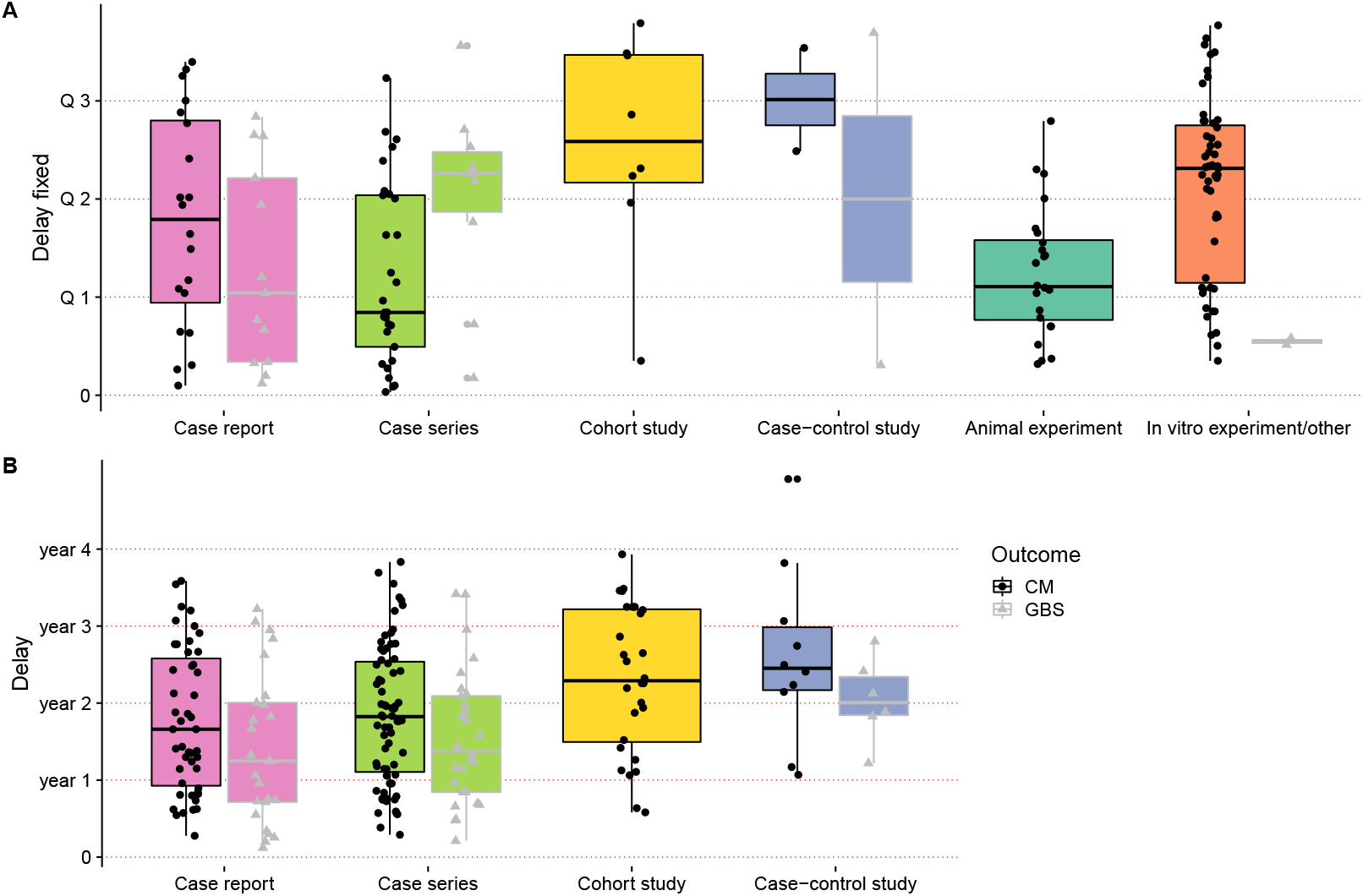
Time to publication, by study type. **A**. The time from the PHEIC declaration (1 February 2016) to the date of publication for studies included up to January 18, 2017. **B**. The time from introduction of ZIKV in the region in which a study was conducted to the date of publication for epidemiological studies included up to January 1, 2019. The box plots show the median an interquartile range, solid black shapes are studies of congenital abnormalities, grey shapes are studies of GBS.

### 3.3 Publication delay

Four out of seven basic research studies published in the first quarter of 2016 appeared as preprints (on www.bioRxiv.com) and had a median publication delay of zero days (Figure 6). The median publication delay increased with time to 107 days (IQR: 66–107 days) by the fourth quarter of 2016. During the same period for epidemiological study designs, the median publication delay increased from 19 days (IQR: 14–22) in the first quarter of 2016 to above 97 days (IQR: 63–138) from the fourth quarter of 2016. In the last quarter of 2018, the publication delay rose to 264 days (IQR: 144–392). On average, the publication delay accounted for 20% (IQR: 11–33%) of the total time from the PHEIC to first publication.

**Figure 6.**
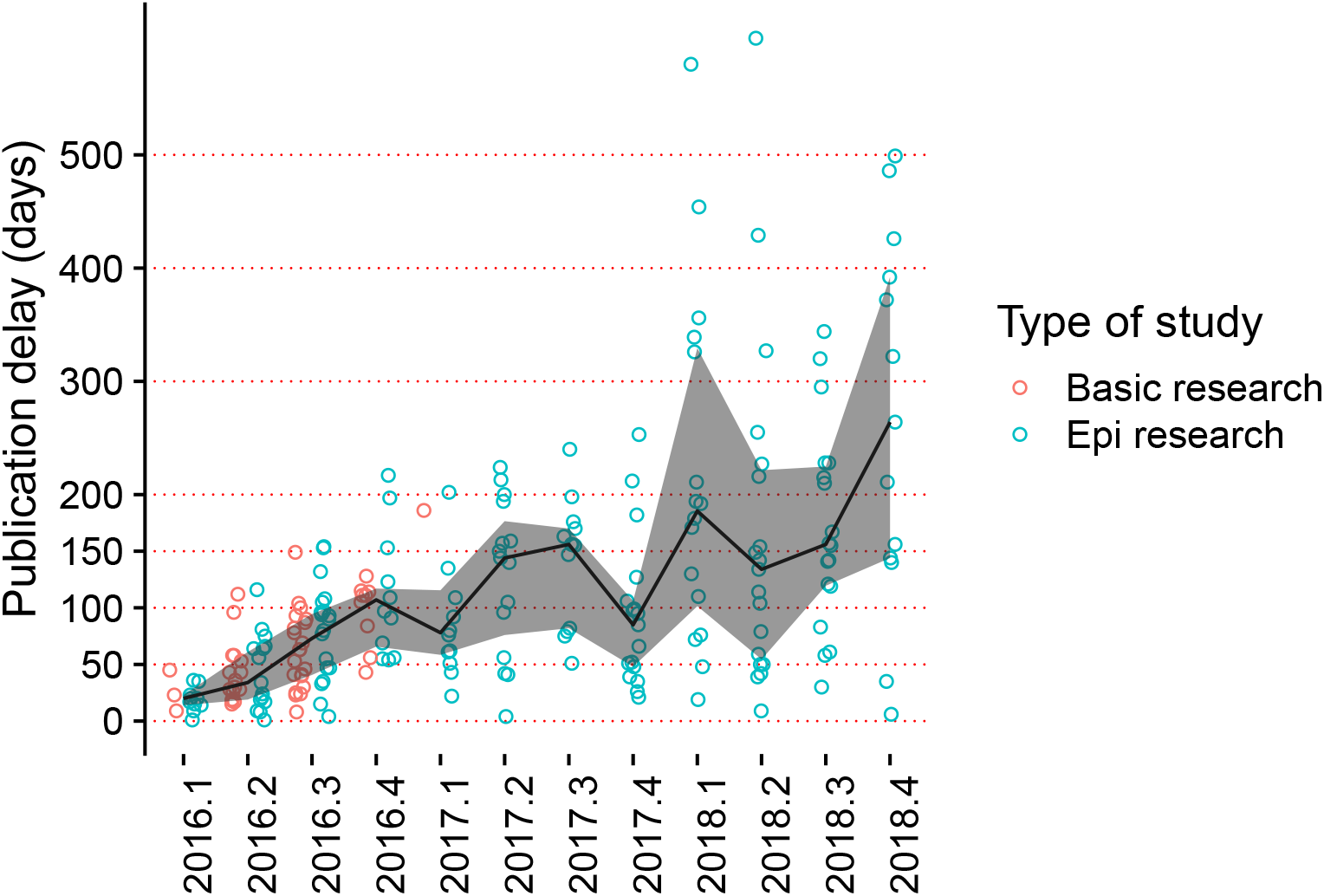
Publication delay, by quarter and by study design. The black line connects the median publication delay (black dots) and the interquartile range (grey ribbon) for all included studies.

## 4 Discussion

In the 2013–2016 ZIKV outbreak, case reports and case series are the first study designs to emerge. Basic research studies appeared rapidly after this. Publication of more robust epidemiological study designs, such as case-control and cohort studies, appeared between 400–700 days after ZIKV was first detected in the region of the study origin. The delay due to the publication process of basic research and epidemiological research was lower at the beginning of the outbreak. A year after the declaration of the PHEIC, the publication delay rose to 150 days. Only a small proportion of publications was available as preprints (16/346).

### 4.1 Strengths and weaknesses of the study

A strength of this study is the pre-specified hypothesis about the time to publication of aetiological research and the use of data from systematic reviews that had screened and selected studies that addressed the causal relationship between ZIKV infection and its adverse outcomes. We calculated additional measures related to the time to publication of research, including delays due to the publication, and thus the time that could have been gained by publishing preprints.

The limited information extracted about each study was a limitation. The time between introduction of ZIKV and the actual publication of a research study is dependent on factors both within and between study designs. There is substantial variation in the time to publication within the study designs. We did not quantify several factors that likely influence this duration such as the size of the outbreak, the research capacity or outbreak preparedness. Small outbreaks or small population sizes limit the opportunity to enrol sufficient patients with adverse outcomes, and unless involved in multi-centre/multi-region studies, these regions are less likely to produce high quality epidemiological studies. The same holds true for regions with limited research capacity, such as appropriate diagnostic facilities and expertise. Outbreak preparedness likely increased over time, with funding increasing after the PHEIC, meaning that initiation of studies started relatively late for regions that were affected earliest by the outbreak. Countries that were affected later in time by ZIKV, might have already had surveillance and diagnostic methodology in place.

The publication delay is a proxy measure, which could not be calculated for all studies; for only 59% (204/346) studies the “received date” was provided. It is unclear whether these data are missing at random. Furthermore, the recorded publication delay could only be calculated for the journal in which a study was published. The true publication delay includes the time taken up by rejection and resubmission. The publication date also ignores dissemination of the findings at conferences or within collaborations. However, here the information is only available to a limited audience. The timing of ZIKV introduction is also a proxy measure, which does not capture the first actual case, but signals the moment at which the health authorities and the research community noted the introduction in that region and thus serves its purpose as a proxy for when research start intensifying. Phylogenetic data suggest that ZIKV was often introduced months before formal detection and notification [18].

### 4.2 Interpretation of the findings

The sequence of emergence of evidence about causality was not exactly as hypothesised (Figure 1). While case reports and case series were the first types of study to be published, findings from animal research were also published quickly. This finding might have been influenced by the more frequent use of preprints to disseminate laboratory research than clinical science [19]. In our study, the time taken to publication of case-control studies and cohort studies was similar, particularly for studies of congenital outcomes. Case-control studies are widely assumed to be quicker to organise and conduct than cohort studies [1, 15]. In the ZIKV outbreak, one case-control study about GBS was published soon after the PHEIC declaration because it used data already collected from the earlier ZIKV outbreak in French Polynesia. An important consideration is the short duration of pregnancy. The cohorts that were fastest to produce results, were cohorts that were already in place for other disease (Dengue, influenza) [20]. Follow up of the outcomes of a disease exposure in pregnancy takes a matter of months. It might therefore not be possible to extrapolate this finding to other conditions in which the outcome takes years to develop.

The rapid, and sustained, publication of a large body of research about ZIKV is a rich resource for meta-research about causality. The declaration of the Ministry of Health of Brazil in November 2015 [5] and the declaration of the PHEIC by WHO in February 2016 [7] catalysed the ZIKV research effort across many different disciplines and resulted in an increase in research funding and a WHO-initiated research agenda [21]. Some of the observed patterns in study design and time publication are influenced by the type of outcome. Investigation of GBS, which is very rare, estimated at 4/10.000 ZIKV infections [22], is likely to be restricted to case-control as cohort studies would take too long to enrol enough participants.

We provide empirical evidence about publication delays during an outbreak of an emerging infection. WHO and others have encouraged rapid dissemination and timely open access to data to help the response during public health emergencies [23]. The ZIKV outbreak and 2013–2016 Ebola virus outbreak in West Africa, emphasised the need for rapid sharing of data. However, the publication delay returned to an average observed in across disciplines within a year after the declaration of the PHEIC; the age of the average preprint before it is published by a journal across different scientific disciplines is 166 days [24]. In medical research, publication of preprints is still underused [19] but the launch of the MedRXiv preprint server (www.medRxiv.org) in June 2019 might signal a change.

### 4.3 Implications for public health, policy and research

Looking back at the ZIKV outbreak and how evidence accumulated on the adverse outcomes will provide guidance for a next outbreak. It provides insight in how evidence accumulates in new causal questions. Specifically for disease outbreaks, we can increase preparedness by the lessons learnt from the Zika virus outbreak. Especially, since disease outbreaks or disease re-emergence continue to happen due to extraneous pressure such as shifts in climate, population growth and increased movement of people either due to displacement or voluntary movement [25].

In a disease outbreak with adverse outcomes that are new or incompletely understood, the full spectrum of evidence needs to be assessed to establish causality. Early in an outbreak, we need anecdotal evidence to drive discovery and explanation [1]. Studies across the different scientific disciplines are informative while we wait for robust epidemiological studies. Here, different frameworks can help us assess the evidence such as Bradford Hill dimensions [11]. We rely on a wide spectrum of evidence in line with how Krieger et al. phrase it: “Robust causal inference instead comprises a complex narrative, created by scientists appraising, from diverse perspectives, different strands of evidence produced by myriad methods.” [26]. Rapid consensus on causality is often needed to form public health guidance.

Not all outbreaks generate the same emerging causal questions, thus deconstructing other causal problems based on study design and timing of evidence will provide more insight in how evidence accumulates. The ZIKV outbreak in the Americas was unique by its size; making rare non-pathognomonic outcomes visible. Also, the 2015–2017 outbreak in the Americas benefited from the outbreak in 2013 in French Polynesia. Much data was collected there, and retrospective analyses confirmed the association between ZIKV infection and adverse outcomes [8]. This resulted in the publication of a case-control study on GBS, rapidly after the declaration of the PHEIC [8]. Likewise, Cauchemez et al. used a modelling approach to estimate the risk of adverse congenital outcomes in French Polynesia retrospectively [27].

### 4.4 Future research

As the SARS-nCoV-2 outbreak continues to unfold, we will apply the same methodology as discussed here to keep track of the accumulation of evidence, the delay in publication and the use of pre-print publishing.

### 4.5 Conclusion

The accumulation of evidence over time in new causal problems seems to follow a hierarchy where case reports and case series were rapidly followed by basic research. During the ZIKV outbreak, robust epidemiological studies, such as case-control studies and cohort studies, took 400–700 days to appear. Causal inference based on a wide spectrum of evidence is therefore essential for early public health guidance in emerging causal problems. Publishing preprint does reduce the delay, and especially in epidemiological research this is an underused tool.

## Data Availability

All data is publicly available.

https://f1000research.com/articles/8-1433

## Notes

### Competing Interest Statement

The authors have declared no competing interest.

### Funding Statement

MJC received salary support from the Swiss National Science Foundation [320030_170069].

